# Efficacy and safety of cyclosporine in the management of coronavirus disease 2019: A protocol for systematic review and meta-analysis

**DOI:** 10.1101/2021.07.03.21259959

**Authors:** Ibtihal Abdallah, Mohamed Aabdien, Mohammed Danjuma

**Affiliations:** Clinical Pharmacy Department, Hamad General Hospital, Hamad Medical Corporation, Doha, Qatar; Community Medicine Training Program, Medical Education, Hamad Medical Corporation, Doha, Qatar; Division of General Internal Medicine, Weill Cornell affliated-Hamad General Hospital, Hamad Medical Corporation, Doha, Qatar

**Author notes:** **Corresponding author:** Ibtihal Mahmoud Abdallah, Clinical Pharmacist, Clinical Pharmacy Department, Hamad General Hospital, Hamad Medical Corporation, Doha, Qatar.

**Keywords:** Cyclosporine, Coronavirus disease 2019, COVID-19, SARS-COV-2, calcineurin inhibitor, ciclosporin

## Abstract

**Introduction:** Cyclosporine may improve the clinical course and outcomes of *Coronavirus disease 2019* (COVID-19) due to its antiviral and anti-cytokine effects as shown in vitro. A few ongoing trials are exploring the benefit of adding it to the standard of care (SOC) of *COVID-19* patients.

**Objectives:** The primary objective is to evaluate the severity of COVID-19, determined by oxygen saturation, intensive care unit (ICU) admission, or the World Health Organization COVID-19 clinical severity scale in patients treated with oral or intravenous cyclosporine added to SOC compared SOC alone or placebo. Secondary objectives include mortality, length of hospitalization, length of ICU stay, and laboratory measurements as well as the safety outcomes of cyclosporine.

**Methodology:** A systematic review and meta-analysis of randomized clinical trials and observational studies that compared cyclosporine to placebo or SOC in COVID-19 patients will be conducted. PubMed, EMBASE, Web of Science, Cochrane Central Register of Controlled Trials, Google Scholar, and ClinicalTrials.gov will be explored for studies that satisfy pre-specified inclusion criteria. Quality assessment of all included studies will be performed. Meta-analyses will be done utilizing random effect models to estimate the effect of cyclosporine on the severity of COVID-19. Heterogeneity will be assessed utilizing Q statistics. The Preferred Reporting Items for Systematic Reviews and Meta-Analyses guidelines will be followed.

**Results:** The result of this synthesis will inform potential changes in the management of COVID-19 patients, especially regarding the role of calcineurin inhibitors. Additionally, it will serve as hypothesis generating for potential future prospective studies.

## Background

### Rational

Coronavirus disease 2019 (COVID-19) is caused by severe acute respiratory syndrome coronavirus 2 (SARS-CoV-2), a member of the family of coronaviruses that infect both humans and animals (1). COVID-19 was declared a public health emergency of international concern by the World Health Organization (WHO) in late January 2020, and it was declared a pandemic as early as March 2020 (2). As of March 2021, SARS-CoV-2 has infected over 100 million individuals around the world, and its related deaths have exceeded two million recorded deaths (3).

Cyclophilin A (CypA) is a protein that belongs to the immunophilins’ family. It is an abundant protein in the cell cytoplasm and has a major role in multiple functions including regulation of gene transcription, immunomodulation, cell signaling, protein folding and trafficking (4). CypA is a putative factor in the pathogenesis of numerous inflammatory conditions, and its expression is increased during such conditions (4,5). Additionally, it facilitates the propagation of several viruses such as Humanimiunodefiency virus type 1 (HIV-1), coronaviruses (CoV), hepatitis B virus (HBV) and hepatitis C virus (HCV). Hence, it is considered a potential attractive target for antiviral therapy (4–6).

Cyclosporine (Cs) is a calcineurin inhibitor (CNI) immunosuppressive drug that is indicated in a wide range of inflammatory and immune conditions, including rheumatoid arthritis, solid organ transplants and nephrotic syndrome amongst others (5). It acts selectively via binding to its cellular receptor, cyclophilin A (CypA), in the cytoplasm of T cells. This binding forms a complex that inhibits calcineurin. Inhibiting calcineurin blocks the translocation of nuclear factor (NF) from the cytosol into the nucleus of the activated T cells. This process inhibits the transcription of genes that encode interleukin-2 (IL-2) and hence prevents the full activation of T cells (5,6).

Calcineurin inhibitors (CNI) have shown some inhibition of viral replication in vitro through the inhibition of CypA (5). Cyclosporine has exhibited inhibition of several coronaviruses in vitro independent from its immunosuppressive effect and at doses and concentrations lower than those needed for immunosuppression (5,7). Evidence suggests that CNIs might have a role in SARS-CoV-2 management. It is speculated that the leading cause of death among COVID-19 patients is acute respiratory distress syndrome, as it is probably associated with a hyperinflammatory state and cytokine-release syndrome (CRS) (8). Hence, drugs that reduce such hyperinflammation through inhibiting various cytokines, recombinant IL-1 blocker anakinra, IL-6 receptor blocker tocilizumab, and corticosteroids are currently part of a cocktail of drug regimen employed in COVID-19 management (8).

Cyclosporine may have a role in mitigating COVID-19 related hyperinflammation through the inhibition of the NF of the activated T cells, and as a result inhibiting IL-2 gene transcription, reducing cell proliferation and the concomitant production of other cytokines (7). A recent cohort study was conducted at the Hospital Universitario Quironsalud Madrid and evaluated the clinical outcomes of tocilizumab, glucocorticoids, lopinavir/ritonavir, hydroxychloroquine, cyclosporine on patients hospitalized for severe COVID-19. Cyclosporie use at a cumulative dose of 300 mg was associated with a significant decrease in mortality (0·24, [0·12 - 0·46], p<0·001) when compared to other therapies and after adjusting for confounding factors such as age, baseline CRP, d-dimer > 2·5 μg/mL, diabetes mellitus, and baseline PaO2/FiO2 (9).

Furthermore, there are a few ongoing randomized clinical trials looking to evaluate the use of cyclosporine for the management of COVID-19 in non-intensive care unit (ICU) admitted patients (10– 12).

### Objectives

This meta-analysis will collate evidence of reported efficacy and safety of cyclosporine in the management of COVID-19.

- Primary objective: To evaluate the severity of COVID-19 as determined by oxygen saturation, intensive care unit (ICU) admission, or WHO COVID-19 clinical severity scale in patients treated with oral or intravenous (IV) cyclosporine ± the standard of care (SOC) compared to patients treated with the SOC alone or placebo.
- Secondary objective: To assess mortality, length of hospitalization, length of ICU stay, and laboratory measurements (D-dimer levels, ferritin, interleukin-6 [IL-6], C-reactive protein [CRP], Lacate dehydrogenase [LDH] and viral load) in patients treated with oral or IV cyclosporine ± SOC compared to patients treated with the SOC alone or placebo.
- Safety objective: To evaluate the safety of cyclosporine in patients treated for COVID-19 through lymphocyte count, serum creatinine/creatinine clearance, and rate of adverse drug reactions.

## Methods

### Review question

*Is cyclosporin use associated with favorable outcome in patients presenting with COVID-19 clinical syndrome?*

### Eligibility criteria

All interventional and observational studies that present primary data (e.g. RCTs, cohort studies, and case series) will be included.

- Population: Adult patients (18 years or older) hospitalized for SARS-CoV-2 infection
- Intervention: Cyclosporine administered via oral or intravenous route
- Comparator: Standard of care or placebo
- Outcome: Primary: Severity score of COVID-19 as per WHO COVID-19 clinical severity scale, oxygen saturation, or rate of ICU admission Secondary: Mortality, number of days of hospitalization, number of days of ICU stay, levels of D-dimer, ferritin, IL-6, CRP, LDH, and viral load Safety: Lymphocyte count, serum creatinine/creatinine clearance, and rate of adverse drug reactions

### Exclusion criteria

- Trials evaluating the use cyclosporine for indications other than the treatment of COVID-19
- Reviews and meta-analysis

### Information sources and literature search

We will search through PubMed, EMBASE, Web of Science, Cochrane Central Register of Controlled Trials, Google Scholar, and ClinicalTrials.gov for published, unpublished, and ongoing studies. We will also search through the bibliographies of relevant articles and contact authors if needed.

We will search through these databases up to September 30, 2021. We will restrict our search to studies done on humans and published in English language.

An example of the search terms that will be used: “Coronavirus disease 2019” OR “COVID-19” OR “SARS-COV-2” AND “cyclosporine” OR “ciclosporin” OR “calcineurin inhibitor”

### Protocol and registration

The protocol is hosted on the International prospective register of systematic reviews PROSPERO (registration number: CRD42020207964)

### Selection process

After initial identification of potential studies to be included in the meta-analysis through the systematic search, the titles will be independently screened and assessed for inclusion by IA and MA after duplicates’ removal. Following that, abstracts of selected studies will be independently evaluated. In case of any uncertainty, the studies will be screened in full text. Finally, full text articles will be critiqued and evaluated for inclusion by IA and MA independently. Study selection will depend on the inclusion criteria. Any disputes will be settled through discussion. The Preferred Reporting Items for Systematic Reviews and Meta-Analyses (PRISMA) guidelines will be followed to guide data extraction and reporting (13).

### Data extraction and management

Extracted data include study information (author name, publication date, study location, sample size, number of events), patient demographics and baseline characteristics (age, gender, weight, comorbidities, concurrent medications), cyclosporine dose and route of administration, and duration. Additionally, comparator’s details will be extracted. Outcome measures will be extracted and categorized into primary, secondary, and safety.

### Risk of bias assessment

The Oxford Quality Scoring System (i.e. Jadad scale) will be used independently by IA and MA to assess the methodological quality of the RCTs included in this meta-analysis. When needed, the Newcastle– Ottawa quality assessment scale (NOS) will be used to assess the quality of observational studies, considering a score of 7 or more of high quality. Funnel plot will be used to assess for publication bias (14,15).

### Data synthesis and analyses

Baseline characteristics of continuous and categorical variables will be reported using means (± standard deviation [SD]), medians (± interquartile range [IQR]), or percentages. The relative risk and 95% confidence intervals (CI) will be used to pool the effect of cyclosporine compared to standard of care or placebo. Heterogeneity of the meta-analysis will be tested with Q statistics, and the extent of inconsistency among studies will be quantified with *I*^*2*^ statistics considering a value of >60% to reflect significant heterogeneity. Random-effect models will be used for this meta-analysis. All statistics will be conducted using *The R* software for statistical computing and graphics (version 4.0.2, Vienna, Austria, 2020) (16).

### Subgroup and sensitivity analyses

Sensitivity analysis will be done to assess the impact of each of the included studies on the point estimate by sequentially deleting each study. Subgroup analysis will be performed if feasible given the nature of available data.

### Patient involvement

The design of this study depends on collection and analysis of existing data. No patients are involved in the proposed study, and no institutional ethical approvals are needed.

## Discussion

We will attempt in this meta-analysis to quantify the effect of cyclosporine from the pool of randomized and observational studies conducted among COVID-19 patients. This review will answer an important clinical question as there is a paucity of evidence regarding therapeutic pharmacological alternatives available, and proven effective and safe, for this patient population (17). Furthermore, most of the currently available options come with a high cost (18). Hence, there is a need to utilize less-costly available treatments should they prove efficacious and safe.

In addition to that, there is an ongoing discussion in the scientific community on whether patients who are already on cyclosporine for other indications, such as solid organ transplant, should continue using it (7). The current practice varies; while some clinicians prefer to discontinue cyclosporine and all immunosuppressants for these patients if they are infected with SARS-CoV-2, others believe that such practice might be withholding effective treatment from these patients (9,19). Hence, there is a need for high quality evidence to inform our current practice, and ongoing RCTs are in place.

We anticipate that this review will be completed and available by December 2021, but this will depend on the completion and availability of results of the ongoing clinical trials. The findings of this review will be disseminated through conferences and peer-reviewed journals.

## Data Availability

This manuscript is a protocol for meta-analysis, hence data is currently unavailable. Data will be made available upon the completion of the meta-analysis

## Contributors

IA and MA developed the study conception, design, and statistical analysis plan. MD reviewed the study protocol and provided critical review of the scientific content and study methodology and analysis plan. All authors read and approved the final manuscript.

## Conflict of Interest

The authors have no conflicts of interest associated with the material presented in this paper.

## Notes

### Competing Interest Statement

The authors have declared no competing interest.

### Funding Statement

No funding is available for this meta-analysis.

### Author Declarations

Ethical IRB approval is not required for this meta-analysis as a synthesis of exiting and available data.

